# Serological surveys in Reunion Island of the first hospitalized patients revealed that long-lived immunoglobulin G antibodies specific against SARS-CoV2 virus are rapidly vanishing in severe cases

**DOI:** 10.1101/2020.05.25.20112623

**Authors:** Dobi Anthony, Frumence Etienne, Lalarizo Rakoto Mahary, Lebeau Grégorie, Vagner Damien, Sandenon Seteyen Anne-Laure, Giry Claude, Septembre-Malaterre Axelle, Jaffar-Bandjee Marie-Christine, Raffray Loïc, Gasque Philippe

## Abstract

Both cellular and humoral immunities are critically important to control COVID19 infection but little is known about the kinetics of those responses and, in particular, in patients who will go on to develop a severe form of the disease over several weeks. We herein report the first set of data of our prospective cohort study of 90 hospitalized cases. Serological surveys were thoroughly performed over 2 month period by assessing IgG and IgM responses by immunofluorescence, immunoblot, Western blot and conventional ELISA using clinical RUN isolates of SARS-CoV-2 immobilized on 96 well plates. While the IgM and, unexpectedly, the IgG responses were readily detected early during the course of the disease (5-7 days post-first symptoms), our results (n=3-5 and over the full dilution set of the plasma 1/200 to 1/12800) demonstrated a significant decrease (over 2.5-fold) of IgG levels in severe (ICU) hospitalized patients (exemplified in patient 1) by WB and ELISA. In contrast, mild non-ICU patients had a steady and yet robust rise in their specific IgG levels against the virus. Interestingly, both responses (IgM and IgG) were initially against the nucleocapsid (50kDa band on the WB) and spreading to other major viral protein S and domains (S1 and S2. In conclusion, serological testing may be helpful for the diagnosis of patients with negative RT-PCR results and for the identification of asymptomatic cases. Moreover, medical care and protections should be maintained particularly for recovered patients (severe cases) who may remain at risk of relapsing or reinfection. Experiments to ascertain T cell responses but although their kinetics overtime are now highly warranted. All in all, these studies will help to delineate the best routes for vaccination.

---

Reunion (RUN) island is one of the outermost region in Europe, part of the Mascareigna archipel and best known for its major epidemic of chikungunya in 2005-2006 affecting over 1/3 of the population (ie. 258 000 cases)^1^ Yet again, the island has been impacted more recently from beginning of March 2020 with the first set of imported cases of the novel coronavirus SARS-CoV2. Both cellular and humoral immunities are critically important to control the infection but little is known about the kinetics of those responses and, in particular, in patients who will go on to develop a severe form of the disease over several weeks. Previous but limited studies have demonstrated that IgG serum levels were higher in severe patients than in non-severe patients two weeks after symptom onset but to what extent this response could last (or not) is unknown.

We herein present the first set of data of our prospective cohort study of hospitalized cases. Serological surveys were thoroughly performed over 2 month period by assessing IgG and IgM responses by immunofluorescence, immunoblot, Western blot and conventional ELISA using clinical RUN isolates of SARS-CoV2 immobilized on 96 well plates.

Among the ninety hospitalized RTPCR+ cases tested at CHU of la Réunion (11 March-20 May 2020), we further fully explored the kinetic of the antibody responses in twenty cases over ten to sixty-four days. While the IgM and, unexpectedly, the IgG responses were readily detected early during the course of the disease (5-7 days post-first symptoms), our results (n=3-5 and over the full dilution set of the plasma 1/200 to 1/12800) demonstrated a significant decrease (over 2.5-fold) of IgG levels in severe (ICU) hospitalized patients (exemplified in patient 1). It has already been demonstrated that seroconversion for IgG and IgM occurred simultaneously and that antibody titers plateaued within 6 days after seroconversion. In contrast, mild non-ICU patients had a steady and yet robust rise in their specific IgG levels against the virus. Interestingly, both responses (IgM and IgG) were initially against the nucleocapsid (50kDa band on the WB) and spreading to other major viral protein S and domains (S1 and S2).

Our original data have several major implications. The techniques used herein clearly argue for the use of conventional methods to perform reliable serological surveys and particularly given the major shortage of commercial kits. Serological testing may be helpful for the diagnosis of suspected patients with negative RT-PCR results and for the identification of asymptomatic infections. Moreover, medical care and protections should be maintained particularly for recovered patients (severe cases) who may remain at risk. Our data suggest that immune protection against SARS-CoV-2 might rapidly vanish over time (in a matter of days) as reported for patients with SARS-CoV-1 (but yet in a matter of months)^2^. RTPCR surveys should be reinforced even in the event of past positive serology. Of further critical note, patient 4 remained SARS-CoV2 + by RTPCR (E and N genes) at day 46. Finally, we believe that the emphasis should be although on studies not only addressing the T cell responses but although their kinetics overtime. All in all, these studies will help to delineate the best routes for vaccination.

Authors thanks all members of staff of clinical boards at the main teaching hospital dealing with COVID patients in Réunion Island. Work is supported by CPER-FEDER COVI-RUN Program to P Gasque.

## Data Availability

All data will be available on request (at).

---

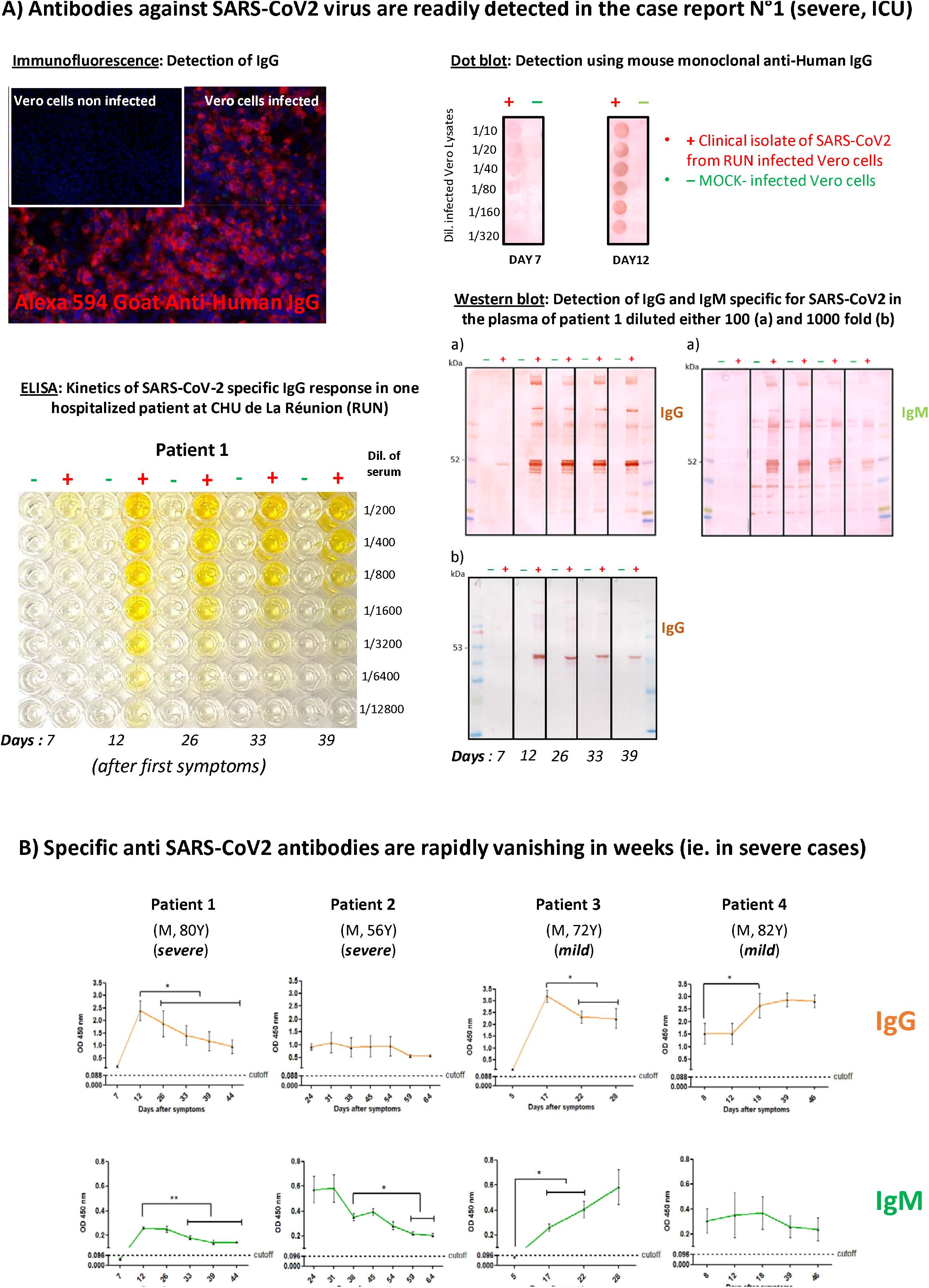

